# Influence of AI-based decision support on shared decision making in hemodialysis: a Wizard of Oz experiment

**DOI:** 10.1101/2023.01.12.23284468

**Authors:** Bilgin Osmanodja, David Samhammer, Roland Roller, Susanne Beck, Aljoscha Burchardt, Wiebke Duettmann, Peter Dabrock, Simon Gerndt, Sebastian Moeller, Simon Ronicke, Klemens Budde

## Abstract

Artificial Intelligence-based decision support systems (AI-DSS) to improve hemodialysis therapy are currently under development. However, the influence of AI-DSS on shared decision making (SDM) in hemodialysis patients has not been studied so far. We performed a Wizard of Oz experiment, using a sham AI-DSS suggesting ultrafiltration volume at the beginning of each dialysis session. We performed 10 patient interviews in 5 patients, and investigated views towards AI, different aspects of the SDM, and the influence of an AI-DSS on SDM in a real-life scenario.

Five main topics were identified: (1) the patient as self-determined, (2) role of the medical staff, (3) other forms of automation, (4) attitude towards AI, (5) needs and preferences for future use of AI. The patients describe novel AI-DSS in dialysis as an opportunity to promote self-determination and to ensure more efficient therapy. At the same time, they describe the special relationship with the nursing staff and physicians, who, from the patients’ point of view, must be given control over an AI-DSS.

This study provides first evidence regarding the influence of AI-DSS on SDM in the context of hemodialysis. Further studies should focus on other aspects that require SDM such as initiation of dialysis, and selection of dialysis modality.

## Introduction

Patients with advanced chronic kidney disease can ultimately require renal replacement therapy (RRT). In Germany, hemodialysis (HD) is the most abundant form of RRT.^1^ For patients and caregivers, this results in a unique treatment setting, where patients are in a healthcare facility 3 times per week, being familiar with the medical staff and routined in their treatment.

From a medical perspective, HD patients frequently experience volume overload, an excess of fluid, which is a major contributor to all-cause mortality and cardiovascular death.^2, 3^ In patients without residual kidney function, fluid accumulates between treatments, and is then removed during the dialysis session. The combination of chronic volume overload and interdialytic fluid accumulation leads to systemic and pulmonary hypertension, arterial stiffness, and left ventricular remodeling.^4, 5^ Additionally, high rates of ultrafiltration required to correct volume overload can lead to complications associated with volume depletion, including systemic hypotension, myocardial stunning and cerebral ischemia.^6–8^.

Determining and maintaining the optimal weight, the so-called dry weight, is difficult even for experienced healthcare providers.^9^ Different approaches are currently studied to improve the fluid management of HD patients, which include biosensors, lung ultrasound, but also machine learning (ML) models.^10^.

Lee et al. developed a recurrent neural network (RNN) based on time-invariant and time-varying features, which is able to predict intradialytic hypotension (IDH) with high accuracy, ranging from AUC-ROC 0.79 - 0.94, depending on the definition of IDH.^11^ Barbieri et al. used an artificial neural network to estimate the minimum systolic blood pressure (SBP), post-dialysis heart rate, weight, and Kt/V, while Chaudhuri et al. predicted relative blood volume decrease of > 6.5% based on optical sensor data.^12, 13^.

These approaches are promising and suggest translation of those ML models into clinically applicable AI-based decision support systems (AI-DSS) in the near future.

But even after such attempts of improving fluid status in HD patients have been thoroughly validated and proven efficacious, AI-DSS have to be embedded into the process of shared decision making (SDM) between patients and healthcare providers.^14–16^ However, little is known about the influence of AI-DSS on SDM in general, and no research has been conducted about this in the specific context of dialysis.

We therefore aimed to study the influence of AI-DSS on SDM in HD patients, using the context of fluid removal during HD for several reasons. (1) As shown above, estimating the amount and rate of fluid removal for a dialysis session is a task that can potentially be modeled by ML in the near future. (2) From our own experience, as well as the literature, we know that while patient involvement into decision making about fluid removal in dialysis is variable, many patients take an active role and even have the final say with respect to ultrafiltration volume.^17^.

(3) Other forms of automation, such as biofeedback systems including blood volume monitoring, are already incorporated in the dialysis treatment.^18^ These can be used to contrast the specific challenges of *AI-based* decision support.

Based on these considerations, the following questions guided our research during conception and conduction:

How are decisions about fluid removal made regularly, and how is it influenced by an AI-DSS making predictions about the optimal fluid removal?

Are current forms of automation, e.g. blood volume monitoring, and *AI-based* decision support perceived differently from patients and physicians and if yes how?

How should an AI-DSS give therapy recommendations and explanations?

## Methods

Instead of studying an experimental AI-DSS like the ones described in the Introduction, we try to shed light on our research questions empirically by performing a Wizard of Oz experiment. This means that patients and physicians are led to believe that an actual AI-DSS is being used to determine the amount of fluid removal, but instead, the system’s suggestions are entered by experienced healthcare personnel that usually would estimate the amount of fluid removal. Hereby, we avoid ethical issues that would arise by using a real AI-DSS to determine a high-risk treatment such as hemodialysis, while still being able to investigate different aspects of human-human-AI interaction in a real-life scenario.

The scenario of hemodialysis treatment offers good conditions for this type of experiment, due to the unique position of the dialysis nurse in the clinical procedure allowing her to “be the AI”.

We included patients receiving maintenance hemodialysis for at least 3 months, to ensure a certain level of patient experience with dialysis and to increase the chance of including patients, who previously have been using other forms of automation in their treatment.

### Standard hemodialysis treatment

Upon arrival, patients are met by the dialysis nurse, accompanied to the scale, and their current weight is measured. Depending on the dry weight that ought to be reached at the end of the dialysis session according to the physician’s prescription, patient-related factors (e.g. current blood pressure, general condition, possible infections, fluid intake during dialysis) and treatment-related factors (e.g. duration of treatment, days elapsed since the last treatment), it is determined how much fluid is to be removed. This usually involves a SDM process between the patient and the dialysis nurse: either the patient is asked how much fluid should be removed or the nurse makes a suggestion based on the dry weight (current weight - dry weight +/- correction factor for fluid intake on dialysis). After considering other factors such as blood pressure, clinical volume overload, and poor general condition, a decision on the target fluid removal is made by the patient and nurse. This is usually close to the patient’s preference and can be corrected by the physician throughout the dialysis session.

After treatment begins, the attending physician visits the patient to discuss medical problems and the settings of the current dialysis therapy, which may include suggestions for changes (e.g. more fluid removal, changes in dialysate composition). In particular, changes in fluid removal are suggested to the patients and, if they agree, the treatment settings are changed.

### Automation using blood volume monitoring

In addition to the dry weight, the patient’s preference, and the clinical assessment by nurses and physicians, automated decision support is already being used to control fluid removal in several HD patients. At our facility, an automated blood volume monitoring system (Hemocontrol biofeedback system, Baxter Deutschland GmbH, Germany) is used in patients with high risk for IDH or high interdialytic fluid accumulation.^18^

Throughout the dialysis session, the system evaluates the blood volume reduction curve and continuously adjusts the ultrafiltration rate and the dialysate conductivity to ensure that the blood volume curve follows a predefined trajectory. This system has been evaluated in clinical trials with variable success in reducing IDH or cardiovascular morbidity^19–23^, but in our experiment helps to contrast *AI-based* against *Non-AI-based* forms of decision support.

### Participant information and consent

Patients and physicians were informed prior to study inclusion that they are participating in a study investigating an AI-DSS for fluid removal in hemodialysis and that the treatment settings are determined by an AI-DSS. The participants were told that the AI-DSS will be used over the course of 3 consecutive dialysis sessions in different settings and patients will be involved to varying degrees in the decision making. The proclaimed aim was to investigate how AI-DSS for HD patients will be used in the future. Patients were also informed that the interaction with the AI-DSS was to be investigated and attitudes, ideas and preferences of patients with respect to AI-DSS were being assessed during semi-structured interviews.

After the study was completed, all subjects (patients and physicians) were informed about the nature of the experiment and told that no AI-DSS was used at any time, but that the treatment data were entered by the nurse to study the human-human-AI interaction and to perform the semistructured interviews in a realistic scenario.

### Study conduction

After arrival, the patients are weighed as usual, their clinical status is assessed (blood pressure, heart rate, temperature, clinical signs of volume overload), and they are asked standard questions about current symptoms, general condition, food intake, and desired fluid removal. This information is entered into a tablet screen **(Figure 1A)**. Subsequently, the target fluid removal as well as the minimum fluid removal necessary and maximum fluid removal possible are determined as clinically indicated by the dialysis nurse **(Figure 1A)** based on the physician’s previous prescription. Afterwards, the treatment is started based on these parameters. Over the course of 3 consecutive dialysis sessions, the exact procedure is slightly changed between 3 different scenarios:

**Figure 1.**
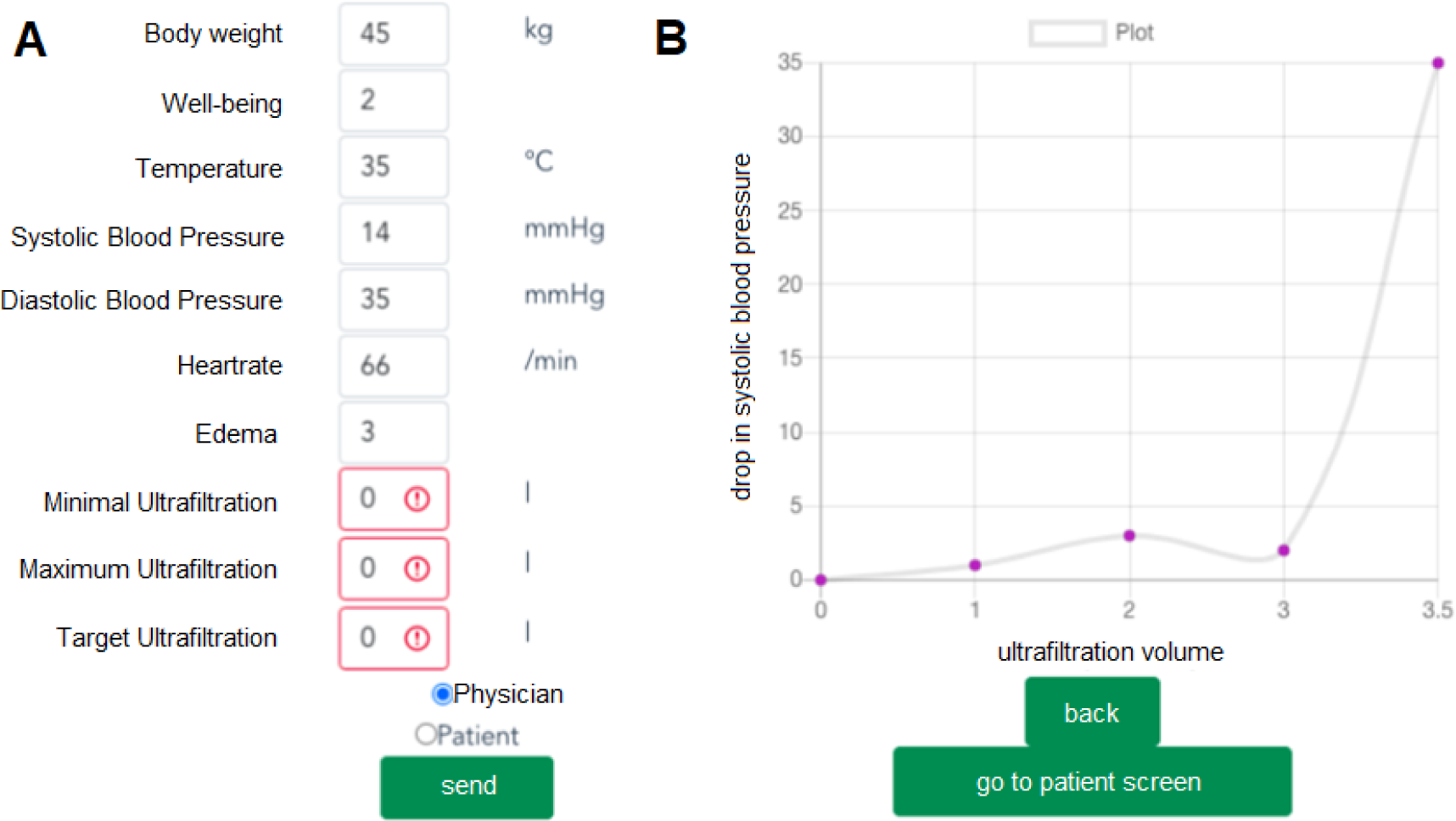
A) Tablet screen designed to enter patient data and clinically indicated fluid removal. **B)** Graphical and textual explanation provided to patients and physicians.

*Scenario 1 (Fully-automated AI):* The sham AI decision is entered into the dialysis machine by the dialysis nurse and the patient is informed afterwards. No SDM occurs, but the treatment begins with the parameters set by the sham AI-DSS.

*Scenario 2 (Conventional AI):* The sham AI decision is communicated to the patient by the dialysis nurse, including both textual and graphical explanation **(Figure 1B).** Then, SDM between patient and nurse occurs.

*Scenario 3 (Value-sensitive AI):* Based on the sham AI decision, two treatment options, based on different patient preference, are provided including a respective textual and graphical explanation:

a. Optimum of long-term dialysis efficiency and treatment comfort. This approach may lead to mild side effects such as cramping in approximately 20% of cases. This corresponds to the maximum ultrafiltration entered by the dialysis nurse.
b. Maximum fluid removal possible without increased risk (<5%) of side effects. This corresponds to the minimum ultrafiltration entered by the dialysis nurse.

Using these suggestions, a shared decision between patient and nurse is made.

During each scenario, the attending physician visits the patients as usual. The graphical and textual explanation generated by the sham AI-DSS are provided to the physician, who can then decide freely how the treatment is continued.

### Semi-structured interviews

In the initial study protocol, semi-structured interviews were planned with patients at the time of study inclusion (baseline interview) and after each dialysis session (follow-up interviews) and for physicians after study inclusion (follow-up interviews). Since the informative content during the follow-up interviews after each dialysis session was limited for the first two patients, we chose to perform only one follow-up interview after completing all three scenarios for each patient thereafter. Physicians were interviewed after performing at least one session with the sham AI-DSS.

During the baseline interviews, the participants were first asked about their assessment of SDM and existing forms of automation for optimizing fluid removal in HD. Then they were asked about their understanding of and their trust in AI-DSS and possible differences to existing forms of decision support such as blood volume monitoring.

During the follow-up interviews, the participants were asked questions about their experience with the AI-DSS and the perceived influence on SDM. A graphical overview of the study course is provided in **Figure 2**. The interviews were recorded and transcribed verbatim.

**Figure 2.**
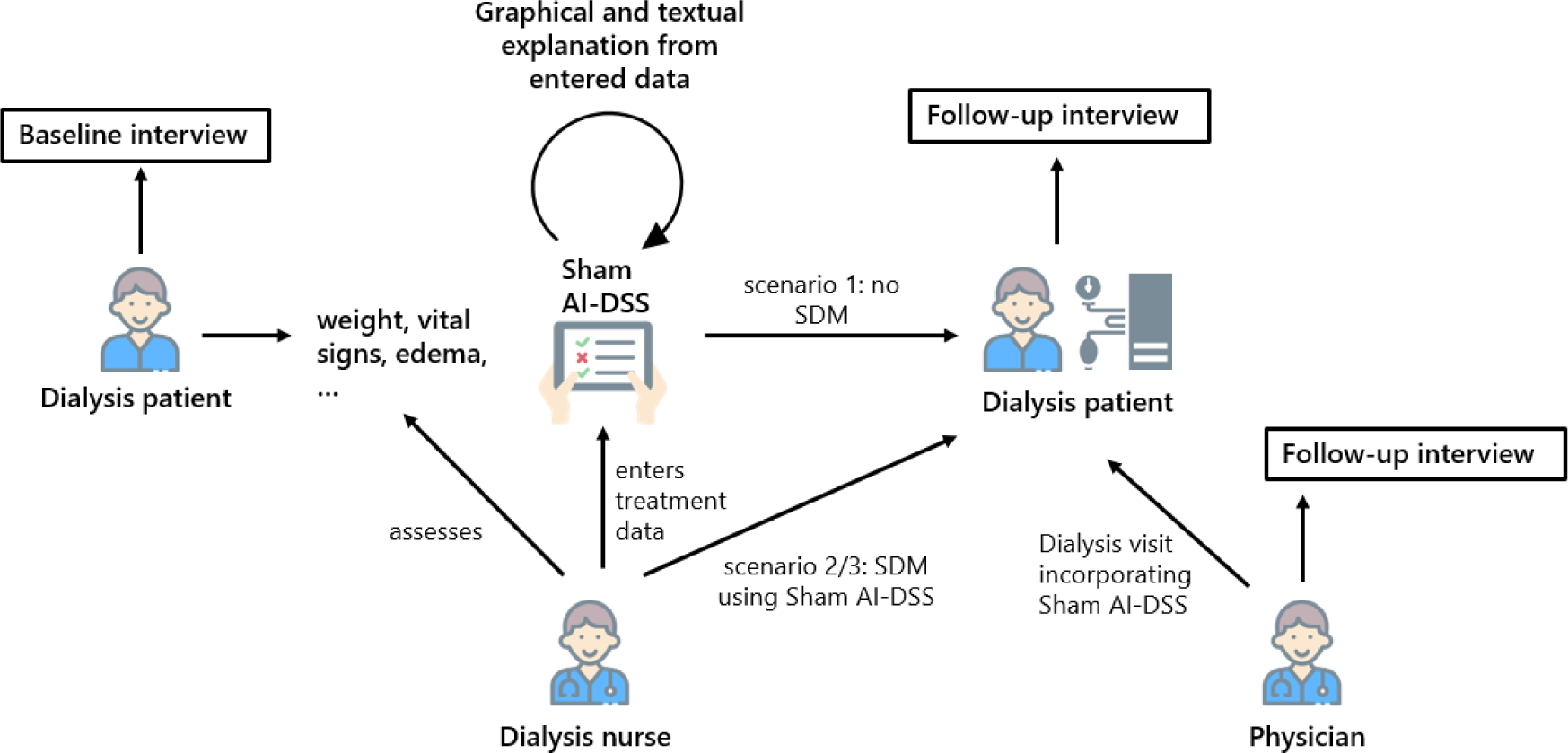
Graphical summary of study course. AI-DSS - artificial intelligence based decision support system. SDM - shared decision making.

### Analysis

The analysis was performed using Atlas.Ti coding program. Two files were created. One for the baseline interviews, the second for the follow-up interviews. For answering the questions raised in the introduction, a separate analysis of the patients’ prior assumptions about hemodialysis treatment and the use of AI from the experience of the experiment seemed suitable. Methodological orientation was provided by Philipp Mayring’s qualitative content analysis ^24^ and certain methodological considerations of grounded theory.^25^ In accordance with qualitative content analysis, the first step was to structure the material. In doing so, we closely followed the topics raised in the questionnaire in order to create a coding scheme. Since it quickly became clear that the patients raised many issues themselves through the open questioning of the semi-structured interviews, we began to use Atlas.Tis function of writing memos and comments to document already first interpretations which was inspired by a common approach of grounded theory.^26^ Finally, the codes, along with the information from the memos and comments, were transferred to excel spreadsheets to discuss the results throughout the research team.

The ethics committee of Charité - Universitätsmedizin Berlin approved this study (EA1/325/21). All participants provided written informed consent.

## Results

From June 2022 until August 2022, five patients, and two physicians completed the study. The median age was 71 years (min: 33 years, max: 81 years), and the median time on dialysis was 2.3 years (min: 1.2, max: 9.4 years). In the patient interviews, five main topics were identified: (1) the patient as self-determined, (2) role of the medical staff, (3) other forms of automation, (4) attitude towards AI, (5) needs and preferences for future use of AI. The content of the interviews is summarized in the following paragraphs, following the structure above. Translations of selected quotes from the interviews are provided in **Table 1**.

**Table 1.** Selected quotes from the patient interviews as well as coding in Atlas.Ti, ordered by category: (1) the patient as self-determined, (2) role of the medical staff, (3) other forms of automation, (4) attitude towards AI, (5) needs and preferences for future use of AI. P - patient, I - interviewer. * added by the author to facilitate the understanding.

| Category | Quote | Codes |
| --- | --- | --- |
| (1) | P: Yes, it always works out the same way. I go to the scale, determine my current weight, compare that with the dry weight, which is actually always quite constant for me over months, and then make a suggestion myself, how many liters we tap. (*P2, BL: 1-1*) | Description of the regular treatment<br>Self-determination |
| (1) | P: So I discuss that, I actually give that as an announcement to the nurse, we draw off so and so many liters. And because it's no problem for me, it's also accepted that way. (.) And I don't really talk to the doctor about the fluid removal when he comes here. (*P2, BL: 1-2*) | Description of the regular treatment<br>Self-determination |
| (1) | P: It's like, I am/ I bring for example, like today, I brought seven, eight hundred milliliters. (.) Now I drink four apple spritzers. So, that's additionally. Then it's 1.6 liters. Then I add a little bit, which kind of | Description of the regular treatment<br>Self-determination |
|  | disappears in the rooms here. Then I'm at 1.7, so that's a good number that's pretty accurate. (P3, BL: 3-31) |  |
| (1) | P: I am // the, I am in the position, because (name) has taught (me*) that, that I set that, my machine, by myself. (.) P: So // I set the temperature, I set the amount of fluid to remove, which I have previously determined at home. Then I already know that and (.) it almost always works. (P3, BL: 3-7) | Description of the regular treatment<br>Current grade of patient involvement |
| (1) | P: No, what I wanted to say. I was going to try food minimally and drink more. (.) Yes. To stay at the same weight, but to drink more than I eat. That's what I have in mind. I haven't tried it yet. Because I noticed that my kidneys always work on Sunday afternoon and Monday morning. It's like they would think, well, probably it doesn't work, we'll have to get back to it. That's what's up. During the week is, (.) I don't know how many milliliters. I could measure it. But when I come from dialysis, I also go to the toilet. But that's maybe, I don't know, ten, twenty milliliters, maybe. Or just ten milliliters. It feels like more. (...) Early always, but that's a maximum of twenty, I think. And under the/ during the day of course now and then a little bit. (P4, BL: 4-22) | Current grade of patient involvement |
| (1) | P: But they've recently, phew, I don't know, two months back, recently been asking the question, how much (fluid*) do we want to draw. They didn't do that before, at the beginning, when I was here. (P4, BL: 4-11) | Decision-making between different actors (patient, nurse, physician) and potential influence of AI |
| (2) | P: No, no. Of course, that's agreed upon but those are, are empirical values and, the people here also know what I can be trusted with. (P1, BL: 1-9) | Description of the regular treatment<br>Decision-making between different actors (patient, nurse, physician) and potential influence of AI |
| (2) | P: They (the physicians*) look at it from time to time, but usually they only say something when I'm not doing so well. Then they check whether the dry weight is still correct, they do an ultrasound of the vena cava or check whether there is water in the legs, blood pressure. But mostly they stay out of it. (P5, BL: 5-10) | Role of the medical staff |
| (2) | P: Um, so all the settings and so on, that's already done by the nurse (.). Yes, actually they do it, I would | Role of the medical staff |
|  | say. The doctor always checks what the settings are, but usually says nothing about it. (P5 BL: 5-11) |  |
| (2) | I get my life here and I'm totally serious about that. [...] I think without dialysis I'm dead. Like this. Consequently, I quite like going here, which is perhaps an important point as well. I feel (.) extremely safe here, in dialysis. When I crawl out of the cab // and am up here and dressed, I feel as safe as the Amen in the church (translated word for word*), because I know that hardly anything can happen to me here. There's always some doctor who can help me. (P3 BL: 3-20,21) | Additional aspects |
| (3) | I // didn't notice the difference (between automated blood volume monitoring and regular dialysis*) (.) I might have felt better a few times, but there are also regularly (treatments*) where I feel better afterwards. (P3 BL: 3-26) | Automated blood volume monitoring |
| (3) | Yes. It is measured continuously, as I understand it, the water content in, in the blood, by, (.) an optical method, I think, by, measuring, the (.) density or something. (P2 BL: 1-11) | Automated blood volume monitoring |
| (3) | I: Did you, if we think about it for a moment, is there anything in common with this Hemocontrol? Were there similarities, differences, anything or did that not really matter to you either? P: (4) No, that has also worked out. (P4 final: 4-3) | Differences between AI and automated blood volume monitoring |
| (3) | P: Well, it seemed to me to be more intelligent than, Hemocontrol, because Hemocontrol is also a rigid program, which took more water from me at the beginning than was good for me. I: Okay, (.) And what was there about/ With this program, what did you experience as more intelligent, so to speak? P: Well, (.) that it didn't come to the blood pressure drop (.) I: So to speak from the/ from the consequence. P: Yes, yes yes. (P2 final: 2-6 / 2-7) | Differences between AI and automated blood volume monitoring |
| (4) | I: Where have you perhaps/ in which/ where has/ where does this (AI*) perhaps still play a role in your life? (...) Can you imagine that somehow? P: (...) I'm thinking about my kitchen appliances right now. But that's also nonsense. (P4 BL: 4-36) | Possibilities of AI<br>Associations with AI |
| (4) | P: Well, we are only at the beginning. Maybe later, when something sensible comes out of what we are trying to do here, that will be/ Just like the device says, hello error, and sounds the alarm. (P4 BL: 4-48) | Possibilities of AI<br>Associations AI and medicine |
| (4) | P: Well, there is always a possibility to determine everything a bit more precisely. Especially the dry | AI in the context of dialysis |
|  | weight is always a bit vague. You can't really determine it exactly. That would be very helpful, of course. *(P5 BL: 5-27)* |  |
| (4) | P: (...) Yes, well, on the basis of the - based on the data that the AI has, I could imagine that the numbers here somewhere become more accurate, if that then first knows more about me, more yes but also very specifically about me, that one fits into my dialysis and that works. I could imagine that well. *(P3 BL: 3-47)* | Possibilities of AI<br>AI in the context of dialysis |
| (4) | Yes, of course. // So I'm still waiting, that this system of, of (that is currently*) a stupid control, at some point will learn to become an intelligent feedback control. [...] For example, the, the potassium withdrawal, that it is continuously measured, and yes, that the machine knows how to proceed. *(P2 BL: 1-28)* | Possibilities of AI<br>AI in the context of dialysis |
| (4) | Yes. I so, just read a few days ago that there are certain rare diseases that doctors can't easily diagnose, but that can be detected by the machine just by taking a picture of the patient's face. And that, I think, is some progress. *(P2 BL: 1-27)* | Possibilities of AI<br>Associations AI and medicine |
| (4) | Well, // with the doctors I've known for a while, when I've gone there regularly, (...) I understand what he means, through his gestures, through his manner, provided he's not having a bad day, he's just/ There was also something like that, when you went home, you thought to yourself: Hey, how did he mean that, hey? Is this really going to start right now (initiation of dialysis*)? Do I have to reschedule everything now? But (.) that was rather rare. But I know, you can almost say, (.) it's like when you come back from a consultation, where a person is sitting, a doctor is sitting or a woman doctor, as when you come back from your parents, into your own apartment. This feeling that comes over is different than when a device tells me something. Yes. And when you're ill, you need something like that. A device like that (.), as nice as it might look afterwards/ No, it wouldn't fit for myself. *(P4 final: 4-9)* | Influence of AI on the roles of the medical staff |
| (5) | I wouldn't call it artificial intelligence, I would call it second intelligence. And the artificial sounds too artificial. And the second one would stand by him (the caregiver). If you were a second man on the trapeze, he holds you. *(P3, final: 3-26)* | Difference of AI and regular treatment |
| (5) | Because she (the nurse*) still makes the settings and I can also give my input, I would say, so clearly, you can accept the suggestions and then of course she |  |
|  | (the AI*) has made her, her contribution to it (.) but I would always leave myself the opportunity to change something again if I say that doesn't quite fit or it's too much or it's too little for the patients. *(P5 final: 5-4)* |  |
| (5) | Yes, so it depends on how deep this informed consent procedure goes. So, I, I don't demand to be informed about the deepest structures of this program, which I might not understand anyway. It would be enough for me if they say, it's happening now with artificial intelligence. And I would wait and see what that does to me. *(P2 BL: 1-36)* | Human control |
| (5) | You could also do patient seminars, but it's just too much of a hassle to go into each room individually. It's nice when there's a synergy, because you have four patients who can talk to each other. And (.) work through the problems. *(P3 final: 3-42)* | Informed consent |
| (5) | Yeah, more variability, for example that intelligence measures how/how long the treatment duration is, whether you could make it longer or shorter, versus the normal course. *(P2 final: 2-4)* | Possibilities of AI |
| (5) | Yes, // it/ there could be more parameters integrated, than just the water content in the blood, no, so also include potassium and whatever else there is. *(P2 final: 2-5)* | Possibilities of AI |
| (5) | P: Yeah, // that would be good if the machine would indicate what's planned, so (.) for example, the/ the course of the/ so if it's about fluid removal, the course will be, with which throughput it starts and/ and how it then continues, and so you could show it as curves here on the screen. *(P2 final: 2-15)* | Preferred explanations |
| (5) | So, if at all // it (the information*) is not important for me, but it is important for the doctor. (.) Because the patient doesn't know all the terms anyway. The patients knows standard things: how much water there is (to be removed*), how much potassium he may eat and so on and that is the most important thing about it. And the rest has to be known by the doctor who sees it, (.) he knows better what is missing. *(P1 final: 1-25)* | Preferred explanations |

### The patient as self-determined

During the baseline interviews, the patients were asked to describe the regular dialysis procedure. Throughout the interviews, all patients describe that they weigh themselves and calculate the difference between their current weight and their dry weight on their own before beginning the treatment *(P2, BL: 1-1)*. If necessary, fluid intake during dialysis, and fluid losses like sweating etc. are considered by the patients *(P3, BL: 3-31)*. The patients are also monitoring themselves at home and use information regarding urine output, fluid and food intake to make more accurate treatment suggestions *(P4, BL: 4-22)*. Most patients consider this process an essential part of treatment, and describe themselves as capable of making self-determined suggestions. One patient even describes programming the dialysis machine by himself, which he learned from the dialysis staff *(P3, BL: 3-7)*. With patients getting more experienced, the nurses are described to ask for their suggestions proactively, which is not the case for less experienced patients *(P4, BL: 4-11).* Based on these suggestions, the final decision is then made together with the medical staff, especially the dialysis nurse. Often, the suggestions by the patient are accepted by the medical staff *(P2, BL: 1-2)*.

### Role of medical staff

Regular dialysis treatment, which often lasts several decades, not only builds a routine, but also a special relationship with the medical staff. Therefore, we asked the patients to describe the interactions and the relationship to the medical staff during the baseline interviews. One patient describes knowledge gained through experience between him and the medical staff. This knowledge would enable medical staff to assess which tasks can be left to the patient’s own discretion and which cannot *(P1, BL: 1-9)*. In addition, the special role of the nursing staff is emphasized. The nursing staff is always described to be present in the immediate environment of the treatment and to carry out regular checkups. If something goes wrong or the patient is unexpectedly unwell, the nursing staff is the first point of contact *(P5, BL: 5-10)*. Physicians, in contrast, tend to be described as passive. While they are involved at the beginning of the treatment, they rarely comment on routine treatment, according to patients *(P5 BL: 5-11)*. However, this passive behavior does not lead to a loss of importance. On the contrary, physicians are given a high level of trust. This is particularly clear from a patient’s statement about his experience of dialysis treatment. He speaks of gratitude and a feeling of security, which is triggered above all by the knowledge that the clinic is an environment in which physicians are ready to help in an emergency *(P3 BL: 3- 20,21)*.

### Other forms of automation

In order to contrast AI-DSS and other forms of automation, we asked the patients about their previous experience with automated blood volume monitoring as one form of automated treatment during the baseline interviews. During the follow-up interviews, the patients were asked to compare the sham AI-DSS with this other form of automation with respect to benefit, understanding, control, and influence on the interaction with the medical staff.

The majority of patients in this study describe having experience with automated blood volume monitoring, but none of them are currently using it in their treatment. The patients describe two reasons for discontinuing this specific form of treatment automation: they either developed episodes of symptomatic hypotension at some point, or did not experience any subjective benefit *(P3 BL: 3-26)*. When asked about the technical details, the understanding is highly variable between different patients. One patient, who happens to be a retired engineer, was able to give a basic explanation of the system’s functional principle *(P2, BL: 1- 11)*, while most patients were able to state the system’s purpose, but not how it worked. While one patient reflected that he should probably know more about such technical aspects, most patients argue that the practical benefit is the most important factor in deciding whether they incorporate such technology in their treatment.

When asked to compare the experimental therapy with the existing form of automation, most patients describe no substantial difference with respect to benefit, control, or interaction with the medical staff *(P4 final: 4-3).* However, one patient described the sham AI system as “more intelligent”, which was attributed to the absence of side effects such as drop in blood pressure *(P2 final: 2-6 / 2-7)*.

### Attitude towards AI

The patients were asked about their expectations towards AI-DSS during the baseline interviews, as well as their experience with the sham AI-DSS during the follow-up interview. What particularly stands out from the interviews is the open attitude of patients towards an increasing and efficient use of data in medicine, even if they report having little contact with comparable technologies in their everyday lives *(P4 BL: 4-36; P4 BL: 4-48).* The hope for dialysis treatment is to be able to determine the correct values even more efficiently and accurately *(P5 BL: 5-27)*. A further step towards personalized treatment is described as a desirable goal (*P3 BL: 3:47)*. AI is considered to have the ability to learn. This specific characteristic is highly valued and seen as an improvement to the previous control options and forms of automation *(P2 BL: 1-28)*. However, from the patients’ perspectives, the potential benefits of AI for medical treatment are not limited to dialysis treatment. For example, one patient reports the potential to detect rare diseases by analyzing just a picture of a person *(P2 BL: 1-27)*. In contrast, the limits of the application of AI-DSS are clarified as well. In particular, one patient explains the advantages of sitting opposite a physician. Human characteristics of a conversation are emphasized. The patient reports that this provides the opportunity of getting to know the physician over time. Even if he is not satisfied with the physician’s decisions at some time, he views such a relationship as necessary to deal with such a serious disease. He could not imagine being treated only by a machine *(P4 final: 4-9)*.

### Needs and preferences for future use

During the follow-up interviews, after completing the three experimental dialysis sessions, the patients were asked to make suggestions about their needs and preferences for future use of AI in the clinic.

The patients report an openness towards technology in general, and AI in particular and emphasize its potential to support caregivers in order to make the treatment safer and more effective. Some patients, by suggesting alternative labels such as “second intelligence” instead of AI, attribute agency to AI-DSS, but also state its subordinate role in decision making *(P3, final: 3-26)*. All patients agree that human agents, namely the patient and medical staff that knows the patient, should control treatment decisions *(P5 final: 5-4)*, and that physicians and the nurses bear the final responsibility for the treatment. The patients argue that the relationship with these agents is fundamental to build trust in their decisions. One patient also attributes responsibility to the organization, since it ultimately decides to incorporate an AI-DSS in the treatment process. When explicitly asked about the responsibility of manufacturers and programmers, the patients attribute some form of responsibility to these agents, but also emphasize that they trust the manufacturing and regulative process to prevent immature systems to be applied in the routine treatment.

Patients were asked if they would warrant additional information to provide informed consent for future A-DSS. Most patients did not demand further technical details, but wanted to be informed about the use of AI in their treatment *(P2 BL: 1-36)*. Another patient suggested that several patients could exchange experiences and their ideas about AI, and maybe should be able to discuss open issues in a patient seminar *(P3 final: 3-42)*.

In the specific context of dialysis, one patient suggested that other parameters such as treatment duration, electrolyte ought to be incorporated into future AI-DSS *(P2 final: 2-4/5).* Another important aspect that was stretched by several patients, is that the graphical representation is valued more important than technical details *(P2 final: 2-15)*. It should inform patients and caregivers regarding the most relevant aspects of the system influence on the treatment. The medical staff should also be provided with additional information necessary to understand the system in greater detail *(P1 final: 1-25)*.

## Discussion

In the present study, we explored patient views on AI-DSS and its influence on SDM in the context of HD by performing a Wizard of Oz experiment, which was accompanied by patient interviews.

In summary, the patients describe novel AI-DSS in dialysis as an opportunity to promote self-determination and to ensure more efficient therapy. At the same time, they describe the special relationship with the nursing staff and physicians, who, from the patients’ point of view, must be given control over an AI-DSS. Thus, the relationship between physicians and patients would hardly change. However, the demands on the medical staff increase, as they must understand the systems in order to adequately inform patients, take over responsibility for the patients treatment, and ensure a trustworthy use of such systems.

We confirmed that hemodialysis patients take over an active role in their regular treatment, which is not limited to the dialysis session, but also includes patient self-monitoring at home, and dietary aspects, which has been reported before.^17^

With respect to SDM, hemodialysis represents a very unique situation, since routine decisions such as the amount of fluid removal have to be made at the beginning of every dialysis session. While we found that caregivers proactively asked for the patients’ suggestions and followed those very closely with respect to fluid removal, we did not study other aspects of the treatment, e.g. dialysis initiation or management of comorbidities. It has been shown for other disease contexts that SDM, while being preferred by physicians, is not as often performed in clinical practice.^27, 28^ It is conceivable that SDM is well implemented for simple decisions in hemodialysis treatment, but informative or paternalistic decision making styles are used for more complex ones.^29, 30^

Regarding the potential use of AI-DSS in hemodialysis, patients identified optimization of dry weight, and fluid removal as one potential use case as well. They also suggested to include other dialysis parameters such as potassium and dialysis duration in order to personalize treatment. When comparing AI-DSS to other forms of automation such as automated blood volume monitoring that are more transparent regarding their functional principle, the patients emphasize the importance of subjective benefit, and reduced to no harm. Technical aspects are less important when deciding whether to use a specific form of decision support or not. This is important, since there is an ongoing debate to what degree transparency is necessary to ensure trustworthy AI.^31^ However, the patients demand some form of information before AI-DSS used in clinical practice. This could be in the form of an informed consent as for other diagnostic or therapeutic procedures. Furthermore, the patients suggest an easy to grasp graphical representation of AI-based decisions or treatment alterations. In the context of dialysis, this means representing changes in fluid removal, and the respective reasons. Using such forms of target-group specific explainability can increase transparency and trust into AI-DSS.^31^

The patients uniformly trust the physicians and nurses to use AI-DSS for their benefit, but also express trust into the regulative process and the manufacturers to prevent non-beneficial or harmful systems being approved at all. This can be interpreted as an invitation to expand current regulations regarding medical devices to account for AI-DSS.

This is the first study to analyze the influence of AI-DSS on SDM in a real-life HD setting by using a Wizard of Oz experiment.

The main limitation of the study is its small sample size, and the selection of long-term dialysis patients. Additional aspects can potentially be found when studying less experienced patients as well. While we studied the routine decision of fluid removal, SDM was mostly studied for other aspects of dialysis therapy before, e.g. choosing the right dialysis modality, the duration and frequency of dialysis so far.^32^ When studying AI-DSS to support such decisions in the future, it is advisable to account for patient values and preferences from the beginning, which can be summarized as value sensitive design.^33^

## Data Availability

All data produced in the present study are available upon reasonable request to the authors.

